# Longitudinal Risk Factors For Developing Depression in Parkinson’s Disease

**DOI:** 10.1101/2021.06.29.21259714

**Authors:** Tarek Antar, Huw R Morris, Faraz Faghri, Hampton Leonard, Mike Nalls, Andrew Singleton, Hirotaka Iwaki

## Abstract

**Background:** Despite the established importance of identifying depression in Parkinson’s disease, our understanding of the factors which place the Parkinson’s disease patient at future risk of depression is limited.

**Methods:** Our sample consisted of 874 patients from two longitudinal cohorts, PPMI and PDBP, with median follow-up durations of 7 and 3 years respectively. Risk factors for depression at baseline were determined using logistic regression. A Cox regression model was then used to identify baseline factors that predisposed the non-depressed patient to develop depressive symptoms that were sustained for at least one year, while adjusting for antidepressant use and cognitive impairment. Common predictors between the two cohorts were identified with a random-effects meta-analysis.

**Results:** We found in our analyses that the majority of baseline non-depressed patients would develop sustained depressive symptoms at least once during the course of the study. Probable REM sleep disorder (pRBD), age, duration of diagnosis, impairment in daily activities, mild constipation, and antidepressant use were among the baseline risk factors for depression in either cohort. Our Cox regression model indicated that pRBD, impairment in daily activities, hyposmia, and mild constipation could serve as longitudinal predictors of sustained depressive symptoms.

**Conclusions:** We identified several potential risk factors to aid physicians in the early detection of depression in Parkinson’s disease patients. Our findings also underline the importance of adjusting for multiple covariates when analyzing risk factors for depression.

## Introduction

Although classified as a movement disorder, recent research has emphasized the challenges to patients and their caregivers that arise from the non-motor symptoms of Parkinson’s disease (PD)[1]. Depression is one such symptom, which is often found to have high comorbidity with PD and has been garnering recognition as one of the most debilitating symptoms to PD patients[2]. Estimates of depression’s prevalence in PD vary from 2.7% to 90%[3], however, meta-analyses have found prevalence across studies to be around 23%[4]. Comorbid depression in PD has been found to be associated with a faster cognitive decline and motor deterioration[5,6]. Additionally, depressive symptoms have been found to account for a large amount of the variance in quality of life scores among PD patients[2].

These findings underlie the importance of detecting depression early to enable clinicians to ameliorate any negative outcomes on patients and their caregivers, whether by initiating antidepressant therapy or offering additional support resources[7]. Previous research has found that younger age, earlier onset of symptoms, female gender, increasing severity of motor symptoms, autonomic symptoms, olfactory deficiency, and a history of psychiatric symptoms are associated with depression during the course of PD[8–11], however, these findings are not consistent across studies[7]. Cross-sectional analyses have also shown that poorer scores on measures of sleep quality and impairment in activities of daily living are also associated with depression[12,13]. However, there is still significant disagreement as to the clinical correlates of depression in PD, and research into risk factors that predict future onset of PD depression is limited[7].

We investigated these variables to test if they were associated with a baseline prevalence of depression using two longitudinal cohorts of Parkinson’s disease - the Parkinson’s Progression Markers Initiative (PPMI) study and the PD Biomarker Program (PDBP).Then we extended our findings to identify factors predicting the development of sustained depressive symptoms in initially non-depressed patients. The use of a Cox regression model to analyze the longitudinal trajectory of depression is rare and provides a unique opportunity to gain greater insight into the progression of PD.

## Methods

### Participants and Measurements

This study utilized data from two longitudinal, clinic-based patient cohorts; PPMI and PDBP. While PPMI was primarily focused on *de novo* patients who had a relatively short duration of diagnosis and were naive to anti-parkinsonian treatment, PDBP had broader inclusion criteria relating to time since diagnosis and medication status. These two cohorts are described in detail elsewhere[14,15].

Patient characteristics such as age and years of education were self-reported at the baseline visit of each study. Other data, such as medication status, were collected prospectively using patient and interviewer completed questionnaires. The Movement Disorder Society Unified Parkinson’s Disease Rating Scale (UPDRS) part III score was used as a measure of motor status, while the UPDRS part II served as a measure of impairment in daily activities. We also used items 2 and 11 of the UPDRS part I questionnaire to screen for baseline hallucinations and constipation respectively [16]. Constipation was screened with a cut-off of ½, representing mild difficulties with constipation, while hallucinations were screened with a cut-off of 0/1, representing any hallucinations. Hyposmia was assesed using the University of Pennsylvania Smell Identification Test (UPSIT)[17] with thresholds normalized for age and sex. Probable REM sleep behavior disorder (pRBD) was screened for using the REM Sleep Disorder Questionnaire with a cutoff of 6 in PPMI[18] and using the first question of the Mayo Sleep Questionnaire in PDBP[19]. The presence of excessive daytime sleepiness was assessed using a cut-off of 9/10 in the Epworth Sleepiness Scale[20,21]. Cognitive impairment was evaluated using the Montreal Cognitive Assessment[22].

Presence of clinically significant depressive symptoms was determined using depression-specific scales from each cohort every 12 months. For PDBP, we used the Hamilton Depression Rating Scale (HDRS)[23] with a cut-off of 9/10[24]. For PPMI, we used the Geriatric Depression Scale Short Form (GDSS)[25] with a cut-off of 4/5[24].

### Data Analysis

A multivariate logistic regression model was used to identify baseline risk factors for depression in each cohort. A dichotomous variable describing the presence of baseline depression was used as the dependent outcome in this model with various continuous and binarized variables as described in Table 2. Predictors were chosen based on a systematic review of potential risk factors associated with PD depression in PubMed using the key terms “depression”, “Parkinson’s”, “longitudinal”, and “risk factors”, returning 80 results most recently on 2/25/21. Any potential factor which was available in both cohorts was assessed for association with depression at baseline. In order to account for fluctuations in depressive symptoms, cases of baseline depression were only included if the patient remained depressed in the following visit. We also implemented a linear mixed-effects model to ascertain whether the screening scores for depression increased over time.

**Table 1.** Baseline Characteristics of Study Participants Subsetted by Presence of Depression at Baseline.

| Cohort | PPMI (N = 418) |  | PDBP (N = 455) |  |
| --- | --- | --- | --- | --- |
|  | Non-Depressed at Baseline (N = 326) | Depressed at Baseline (N = 92) | Non-Depressed at Baseline (N = 388) | Depressed at Baseline (N = 67) |
| **<u>Age</u>** |  |  |  |  |
| - Mean (SD) | 62.3 (9.22) | 59.5 (11.1) | 64.8 (8.88) | 66.0 (9.62) |
| **<u>Disease Duration</u>** |  |  |  |  |
| - Median [Q1, Q3] | 0.36 [0.21, 0.67] | 0.35 [0.22, 0.75] | 3.64 [1.42, 6.70] | 7.10 [3.56, 13.4] |
| **<u>Follow-Up Time</u>** |  |  |  |  |
| - Median [Q1, Q3] | 6.75 [5.08, 7.67] | 6.75 [4.90, 7.23] | 3.00 [2.50, 3.50] | 3.00 [1.50, 3.00] |
| **<u>Race</u>** |  |  |  |  |
| - White | 299 (91.7%) | 81 (88.0%) | 366 (94.1%) | 61 (91.0%) |
| - Non-White | 27 (8.3%) | 11 (12.0%) | 25 (6.4%) | 4 (10.5%) |
| **<u>Gender</u>** |  |  |  |  |
| - Male | 210 (64.4%) | 64 (69.6%) | 235 (60.4%) | 36 (53.7%) |
| - Female | 116 (35.6%) | 28 (30.4%) | 154 (39.6%) | 31 (46.3%) |
| **<u>Years of Education</u>** |  |  |  |  |
| - Mean (SD) | 15.7 (2.86) | 15.0 (2.97) | 16.1 (2.47) | 15.4 (2.46) |
| **<u>UPDRS Part II Total</u>** |  |  |  |  |
| - Median [Q1, Q3] | 6.00 [3.00, 9.00] | 7.00 [4.00, 10.25] | 7.00 [3.00, 12.00] | 14.0 [9.50, 22.00] |
| **<u>UPDRS Part III Total</u>** |  |  |  |  |
| - Median [Q1, Q3] | 6.00 [14.00, 26.00] | 7.00 [15.00, 26.00] | 18.00 [12.00, 29.00] | 28.00 [17.50, 49.00] |
| **<u>MOCA Total Score</u>** |  |  |  |  |
| - Median [Q1, Q3] | 28.00 [26.00, 29.00] | 27.50 [26.00, 29.00] | 27.0 [25.00, 28.0] | 25.0 [22.00, 28.00] |
| **<u>REM Behavior Disorder</u>** | 75 (23.0%) | 39 (42.4%) | 165 (42.4%) | 37 (55.2%) |
| **<u>Excessive Daytime Sleepiness</u>** | 26 (8.0%) | 10 (10.9%) | 62 (15.9%) | 18 (26.9%) |
| **<u>Hyposmia</u>** | 215 (66.0%) | 69 (75.0%) | 284 (73.2%) | 55 (82.1%) |
| **<u>Constipation</u>** | 104 (31.9%) | 34 (37.0%) | 195 (50.1%) | 44 (65.7%) |
| **<u>Hallucinations</u>** | 6 (1.8%) | 6 (6.5%) | 34 (8.7%) | 18 (26.9%) |
| **<u>On Antidepressants</u>** | 27 (8.3%) | 11 (12.0%) | 74 (19.0%) | 29 (43.3%) |
| **<u>On Antiparkinson Medication</u>** | 0 (0.0%) | 0 (0.0%) | 357 (91.8%) | 63 (94.0%) |

Only baseline non-depressed patients from each cohort were included in the subsequent analysis

**Table 2.**
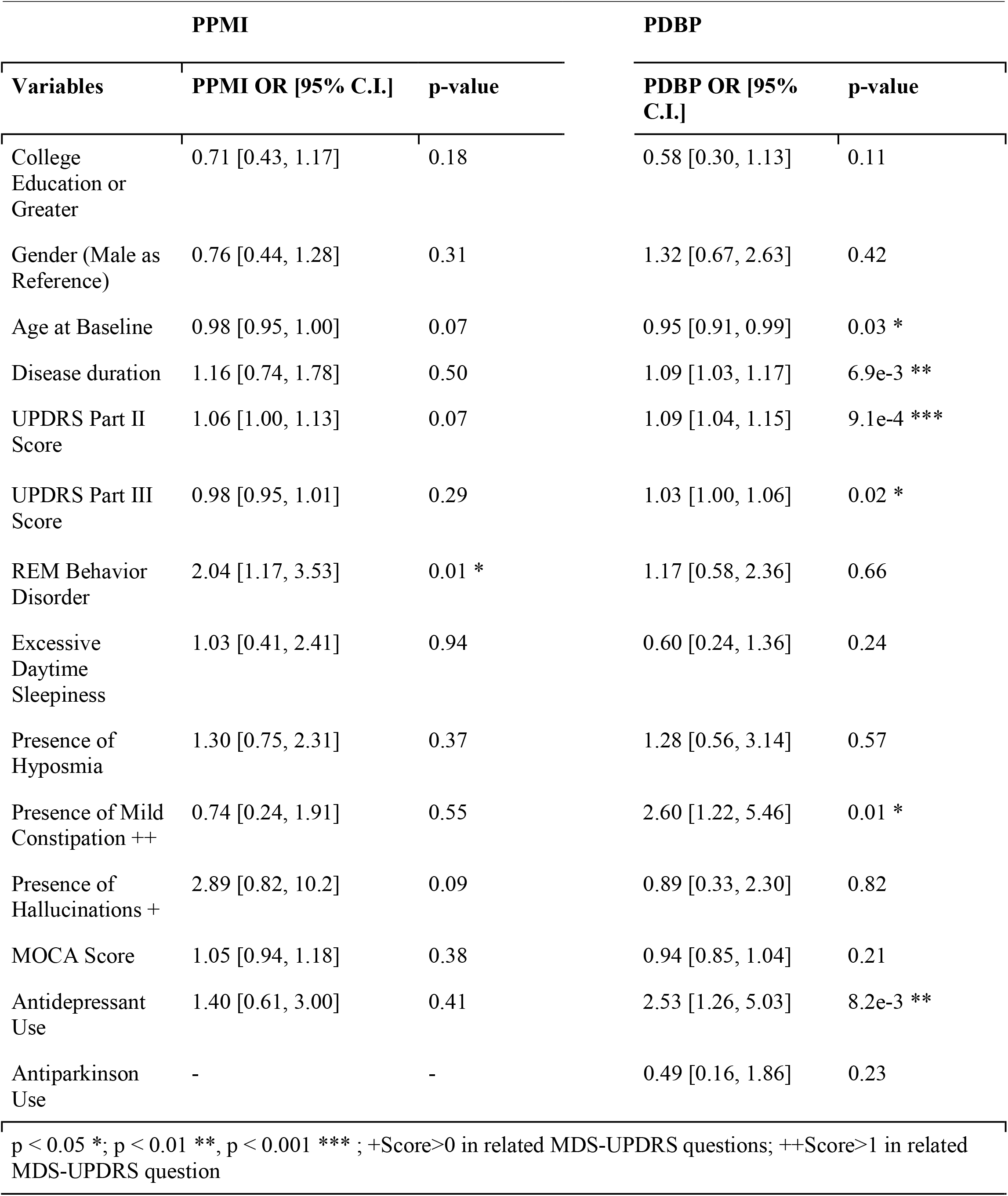
Summary of the Logistic Regression Analysis for the Baseline Depression.

| PPMI |  |  | PDBP |  |
| --- | --- | --- | --- | --- |
| Variables | PPMI OR [95% C.I.] | p-value | PDBP OR [95% C.I.] | p-value |
| College Education or Greater | 0.71 [0.43, 1.17] | 0.18 | 0.58 [0.30, 1.13] | 0.11 |
| Gender (Male as Reference) | 0.76 [0.44, 1.28] | 0.31 | 1.32 [0.67, 2.63] | 0.42 |
| Age at Baseline | 0.98 [0.95, 1.00] | 0.07 | 0.95 [0.91, 0.99] | 0.03 * |
| Disease duration | 1.16 [0.74, 1.78] | 0.50 | 1.09 [1.03, 1.17] | 6.9e-3 ** |
| UPDRS Part II Score | 1.06 [1.00, 1.13] | 0.07 | 1.09 [1.04, 1.15] | 9.1e-4 *** |
| UPDRS Part III Score | 0.98 [0.95, 1.01] | 0.29 | 1.03 [1.00, 1.06] | 0.02 * |
| REM Behavior Disorder | 2.04 [1.17, 3.53] | 0.01 * | 1.17 [0.58, 2.36] | 0.66 |
| Excessive Daytime Sleepiness | 1.03 [0.41, 2.41] | 0.94 | 0.60 [0.24, 1.36] | 0.24 |
| Presence of Hyposmia | 1.30 [0.75, 2.31] | 0.37 | 1.28 [0.56, 3.14] | 0.57 |
| Presence of Mild Constipation ++ | 0.74 [0.24, 1.91] | 0.55 | 2.60 [1.22, 5.46] | 0.01 * |
| Presence of Hallucinations + | 2.89 [0.82, 10.2] | 0.09 | 0.89 [0.33, 2.30] | 0.82 |
| MOCA Score | 1.05 [0.94, 1.18] | 0.38 | 0.94 [0.85, 1.04] | 0.21 |
| Antidepressant Use | 1.40 [0.61, 3.00] | 0.41 | 2.53 [1.26, 5.03] | 8.2e-3 ** |
| Antiparkinson Use | - | - | 0.49 [0.16, 1.86] | 0.23 |

p < 0.05 \*; p < 0.01 \*\*, p < 0.001 \*\*\* ; +Score>0 in related MDS-UPDRS questions; ++Score>1 in related MDS-UPDRS question

Prior to time-to-event analysis, the data was subset so that the sample analyzed included only patients who did not have depression at baseline. The demographics of this subset are described in Table 1. We found it most effective to dichotomize years of education based on a cut-off of 16 years, corresponding to at least a college education in the United States. In order to account for patients who only experienced limited episodes of depression, events were only preserved if the patient met the criteria for depression in the visit following their initial onset. Thus, a depression event was defined as one where symptoms lasted for a period of at least one year. As a result, patients who were only present for a single visit were removed and depression events in the last visit were not included in the analysis. Cases were censored at the last visit recorded or with the onset of depression as defined above.

A cox model was then used to identify baseline factors that placed a PD patient at risk of developing depression. Predictors for this model were chosen based on whether a given factor was significant in the logistic regression for either cohort. Additionally, time-dependent covariates for MoCA score and antidepressant status were included to adjust for the known effects that cognitive deterioration and antidepressants have on depressive symptoms in PD[7]. Antidepressant status also functioned as a marker of clinically diagnosed depression, which we were unable to assess in PDBP due to the lack of medical history data. A meta-analysis of the Cox model results was then conducted using a random-effects model with inverse-variance weighting. Significant heterogeneity was determined by an I^2^ of greater than 75%. All statistical analyses were carried out in R version 4.0.3. Analysis scripts are available at https://github.com/GP2code/LongPDDepRisk. The significance of variables was determined using an alpha of 0.05 in a two-sided test.

### Ethics

Participants’ information was obtained under appropriate written consent and with local institutional and ethical approval. The study protocols were approved at the local institutional review boards, and the participants provided written informed consent.

## Results

Detailed demographics and baseline clinical variables are described in Table 1. Notable differences included the percent of patients on various medications and duration of diagnosis, which is due to PPMI primarily recruiting de novo PD patients. Another notable difference was that the median follow-up for PPMI was longer than in PDBP (7 years vs 3 years).

Table 2 shows the results of our logistic regression analysis at baseline. In PPMI, the only risk factor for depression was pRBD. The recruitment approach of PPMI necessitated medication-naive patients, as such we were unable to assess antiparkinson medication’s association with depression at baseline. In PDBP, younger age, a greater duration of diagnosis, greater severity of motor symptoms, increased difficulty with daily activities, antidepressant use, and at least mild difficulties with constipation were all independently associated with depression.

Figure 1 and Supplemental Table 1 illustrate that depression became increasingly common with disease progression in both cohorts. Figure 1 shows that using any measure of depression, the majority of patients developed clinically significant depressive symptoms during the course of each study. Supplemental figure 1 illustrates this relationship using the additional UPDRSd measure of depression. Across all measures of depression, there was a significant positive association between time since baseline and scores on depression screening inventories (Supplemental Table 2).

**Figure 1.**
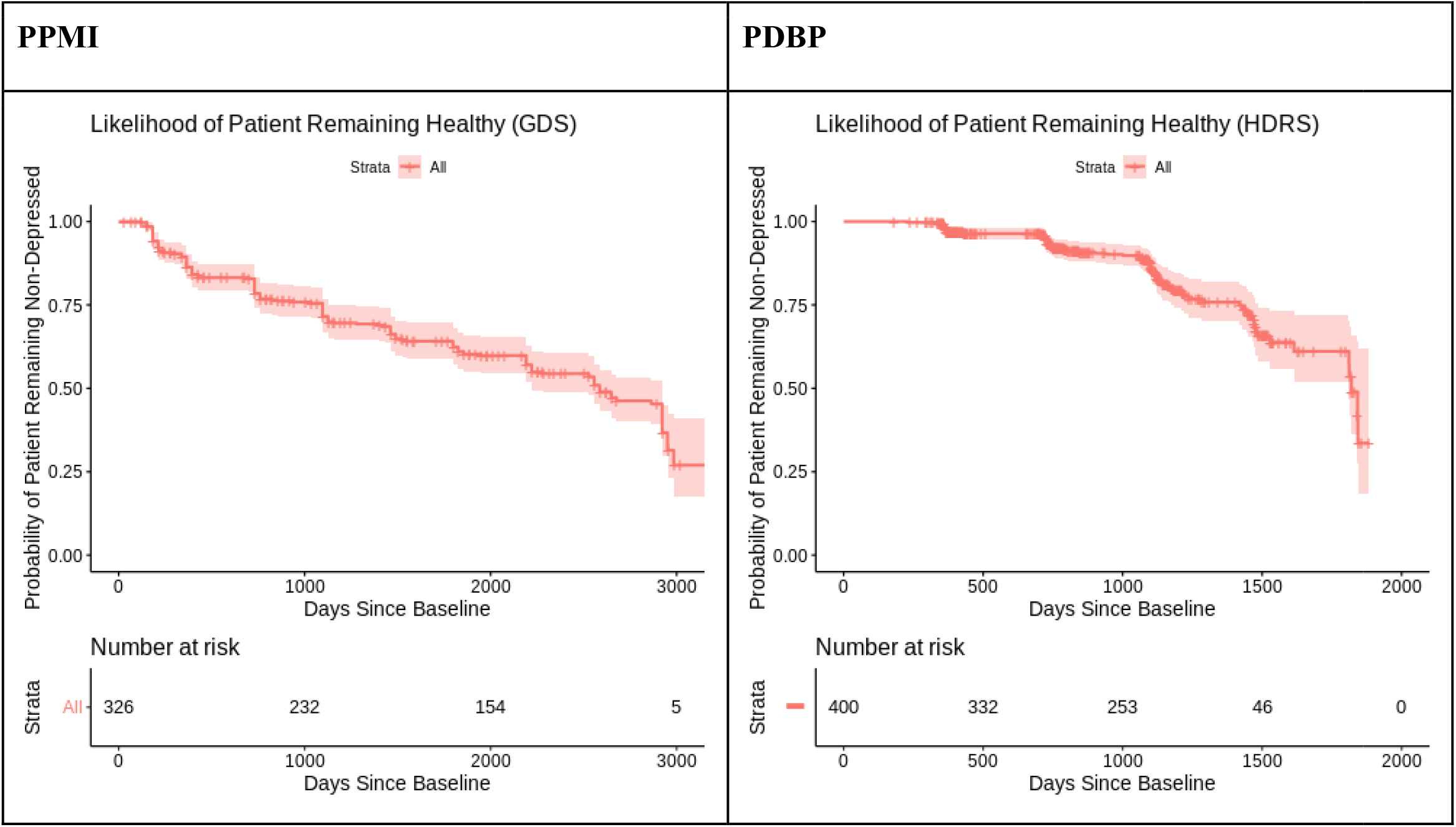
Kaplan-Meier curve showing the probability that an initially non-depressed patient will remain non-depressed, illustrating depression as a progressive symptom of PD.

The different appearance in the two Kaplan-Meier (KM) curves was likely due to baseline differences of disease durations, as when we shifted the KM curve in PPMI’s baseline to year 3 (the mean difference of disease duration), the two KM curves were similar. (Supplemental Figure 2).

Table 3 shows the results of cohort-level analysis results of the multivariable Cox models and a random-effects meta-analysis. In the Cox model, we adjusted for MoCA score and antidepressant status at each visit. There were three significant baseline predictors for the development of depression across the cohorts; pRBD, UPDRS part II score, and hyposmia. There was heterogeneity between cohorts with regard to constipation, but not with any of the other predictors.

**Table 3.**
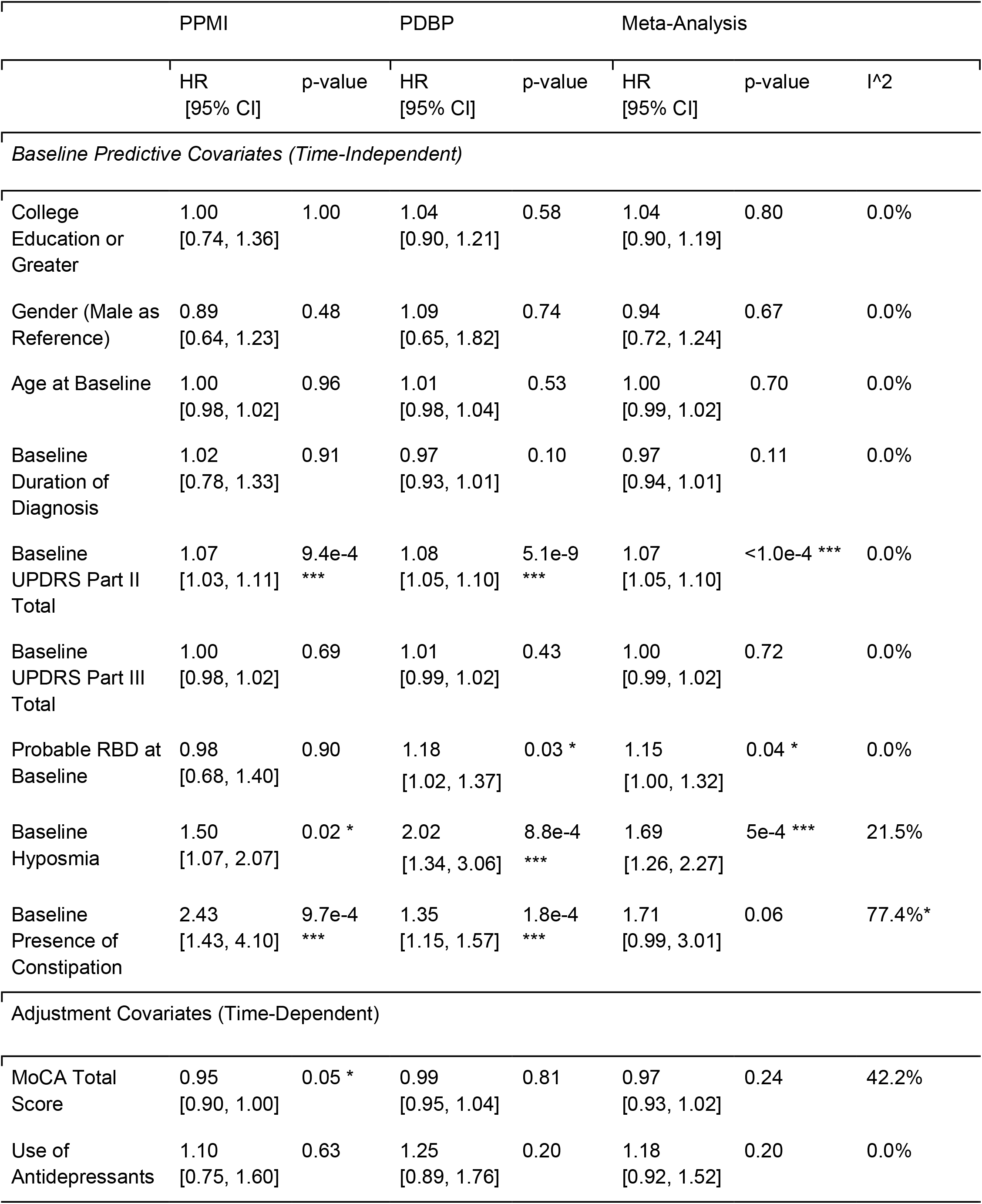
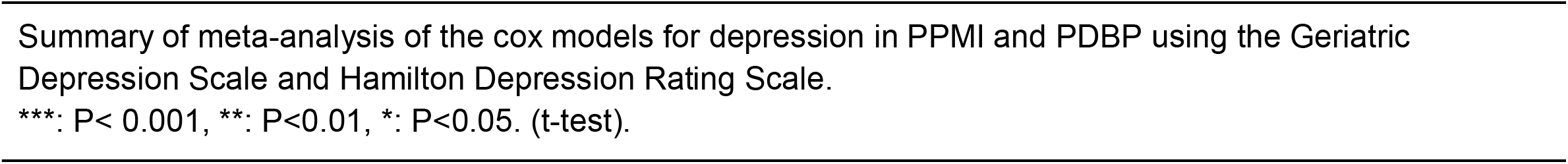
Meta-analysis of Cox Regression Models for Depression.

## Discussion

Given the widespread association of depression with a variety of clinical outcomes, the importance of identifying high-risk patients early on cannot be overstated. We analyzed longitudinal data from 874 participants and implemented a cox regression model which established that a patient’s degree of impairment in activities of daily living and the presence of a probable REM sleep disorder (pRBD) can both be used to predict the onset of depression in non-depressed patients. We also found that pRBD, age, diagnosis duration, impairment in daily activities, severity of motor symptoms, mild constipation, and antidepressant use are among the baseline risk factors for depression. Moreover, we identified heterogeneity in depression risk associated with constipation, indicating potential differences in depression between different stages of Parkinson’s. This emphasizes the need for screening instruments that can identify at-risk patients based on the constellation of their symptoms and clinical variables, as well as the stage of their disease. Our study represents only a first step in the creation of such instruments and further research into the predictive factors of depression in PD is required. It is worth emphasizing that we adjusted for MoCA score, thus accounting for any confounding effect that cognitive decline may have had on the onset of depression. We also included antidepressant status as a time-dependent covariate, which functions as the best available marker of a history of depression at baseline, as well as a marker of physician-diagnosed depression as the study progressed.

Among the covariates that were significant longitudinal predictors, UPDRS part II score is perhaps the most subjective measure of a patient’s symptoms. Rather than assessing any individual symptom, the UPDRS part II assesses a patient’s experience of living with Parkinson’s disease by focusing on their constellation of symptoms. Interestingly, UPDRS part III score was not related with future depression, even when we removed UPDRS part II score from the model. This may indicate that UPDRS part II’s predictive ability comes not from a quantification of motor symptom severity, but from the patient’s more subjective appraisal of their symptom severity. From a clinical perspective, it is also understandable that individuals who report a greater baseline difficulty with their daily activities will likely face mounting difficulties as their disease progresses. There is also a potential effect from recall bias, as an individual with a poorer perception of health status will likely report greater difficulty in their daily activities and be more likely to develop depressive symptoms. Prior research has also established that depression exerts an indirect effect on the quality of life via activities of daily living[26], offering another possible explanation for this finding. Self-report measures such as the UPDRS Part II are also subject to recall bias. Furthermore, the UPDRS part II score was only associated with depression at baseline in PDBP, but was associated with future depressive symptoms in both cohorts. Given PPMI’s status as a *de novo* cohort, this would indicate that impairment in activities of daily living is an especially important factor as Parkinson’s disease progresses.

The remaining longitudinal risk factors, pRBD, hyposmia and constipation, have been identified as symptoms that may precede the onset of Parkinson’s by several years. The potential mechanism for their status as prodromal symptoms is thought to be due to the olfactory bulb, vagus nerve, and brainstem as being induction sites for Lewy pathology before it begins moving into the substantia nigra and cortex[27]. Given these factors’ longitudinal association with depression, this may provide support for depression being a primary symptom related to the physiological progression of the disease, rather than a secondary symptom developing as a response to worsening symptoms. However, despite there being ample research on these symptoms before the onset of motor symptoms, less is known about how these symptoms can be used to understand the progression of Parkinson’s disease[10,11]. Previous studies have associated pRBDs with PD depression in a cross-sectional manner[28], however, methodological limitations prevent the isolation of this effect from the confounding effects of cognitive and functional deficits. Including measures of functional and cognitive status in our models allows us to draw more meaningful conclusions about the effects of pRBD. With regard to hyposmia, there are cross-sectional findings supporting a relationship between olfactory dysfunction and non-motor severity in both the prodromal and symptomatic phases of Parkinson’s [10], however our study extends these findings to establish hyposmia as a predictive factor specifically towards the future development of depression. Constipation was a significant predictor in both cohorts, however it was not significant in the meta-analysis, likely due to heterogeneity between cohorts. Given that the main sources of heterogeneity between the two cohorts were medication status and disease duration, it is likely that one of these factors is responsible for this heterogeneity. Thus, it is possible that constipation is a stronger predictor either earlier in the course of Parkinson’s disease or while the patient is naive to antiparkinson medication or a mixture of the two. Regardless, this finding may point to potential heterogeneity in predictive factors for depression in the early and late stages of Parkinson’s and underlies the need for further research into symptoms of autonomic dysfunction.

The PPMI and PDBP cohorts do represent different subsets of the population of individuals with Parkinson’s disease, so there is some limitation in our ability to harmonize data between the two. For example, there was a notably larger proportion of PPMI patients who had depression at baseline. This may indicate that people with PD are less likely to join a study later in their disease course if they are suffering from depression. There was also not a single baseline risk factor that was common between the two cohorts. Initially, we suspected this may be due to heterogeneity between the two cohorts. However, our meta-analysis found no evidence of heterogeneity between any covariates except for constipation, indicating that our risk factors are similarly associated with depression throughout the course of the disease. This does not exclude the possibility that certain factors exhibit stronger associations at different stages of the disease, but these findings ultimately provide only putative indications of there being variation between the early and late stages of PD. The possibility of there being variation in depression risk factors between the early and late stages of PD represents an interesting topic for further study, especially since it may provide some explanation as to the disagreement in prevalence and risk factors between studies. Additionally, there were several measures which differed between our two cohorts. The most significant was the difference in depression-specific scales; the GDSS was used in PPMI while the HDRS was used in PDBP. Prior studies have raised concerns about there only being a moderate correlation between the GDSS and HDRS[29], however other analyses of the two scales have found they both exhibit good sensitivity and specificity for the diagnosis of depression in PD[24]. The presence of pRBD was also assessed with different methods, however both have been found to have good sensitivity and specificity for screening purposes[30]. Our longitudinal analysis model was also limited in that it only accounts for the first onset of an event, thus we can’t identify which patients will remain depressed and how their risk factors may differ. We did attempt to mitigate this limitation by only preserving depression events that were sustained for at least a year and we did find that overall depressive symptoms did increase over time.

Our study also had several strengths which are worth considering. A strength of our baseline analysis relative to other studies was the adjustment for a variety of demographic and clinical variables. This is especially important when analyzing outcomes with multifaceted causes such as depression, as many of its risk factors are interrelated, and examining one in isolation often fails to paint a complete picture. The longitudinal model allows for a robust method of identifying associations with future depression. This is particularly important to identifying at-risk patients early in their disease so they can receive timely management of their symptoms. Despite differences in measurement and populations between the two cohorts, we still had significant results without heterogeneity, pointing to the generalizability of our findings.

## Data Availability

Data from the Parkinson's Progression Markers Initiative (PPMI) was obtained through the Image & Data Archive (https://ida.loni.usc.edu/). Data from the Parkinson's Disease Biomarkers Program (PDBP) was obtained from the Global Parkinson's Genetics Program (https://parkinsonsroadmap.org/gp2/). Analysis scripts are available at https://github.com/GP2code/LongPDDepRisk. 

## Acknowledgments

We thank all study participants and their families, investigators, and anyone else involved in the Parkinson’s Progression Markers Initiative (PPMI) study and the Parkinson’s Disease Biomarker Program (PDBP).

PPMI—a public-private partnership—is funded by The Michael J. Fox Foundation for Parkinson’s Research and funding partners, including AbbVie, Allergan, Avid Radiopharmaceuticals, Biogen, BioLegend, Bristol-Myers Squibb, Celgene, Denali Incorporated, GE Healthcare, Genentech, GlaxoSmithKline, Eli Lilly and Company, Lundbeck, Merck & Co., MesoScale Discovery, Pfizer, Piramal, Prevail Therapeutics, Roche, Sanofi Genzyme, Servier Laboratories, Takeda, Teva, UCB, Verily, Voyager Therapeutics, and Golub Capital (www.ppmi-info.org/fundingpartners). We would like to thank the Parkinson’s community for participating in this study to make this research possible.

This work was supported in part by the Intramural Research Program of the National Institute on Aging, National Institutes of Health, part of the Department of Health and Human Services (project ZO1 AG000949).

## Notes

### Competing Interest Statement

 Tarek Antar received a Postbaccalaureate Intramural Research Training Award from National Institute Aging. Andrew Singleton has received grants from the Michael J Fox Foundation for Parkinson's Research, the Department of Defense NETPR Program (Grant IAA-XAG16001- 001-00000), and the Aligning Science Across Parkinson's Initiative. The authors otherwise have no financial interests to report.  

