## Supplemental Figures and Tables for "Longitudinal Risk Factors For Developing Depression in Parkinson’s Disease"

**Supplemental Figure 1**

Kaplan-Meier curves with number at risk tables for comparison of the depression measures used in both cohorts.

| **PPMI** | **PDBP** |
|---|---|

**A**

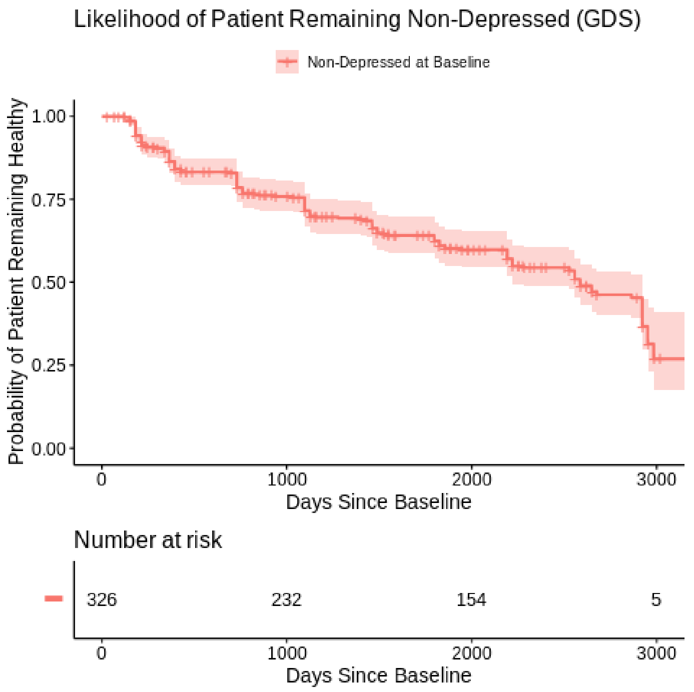


**B**

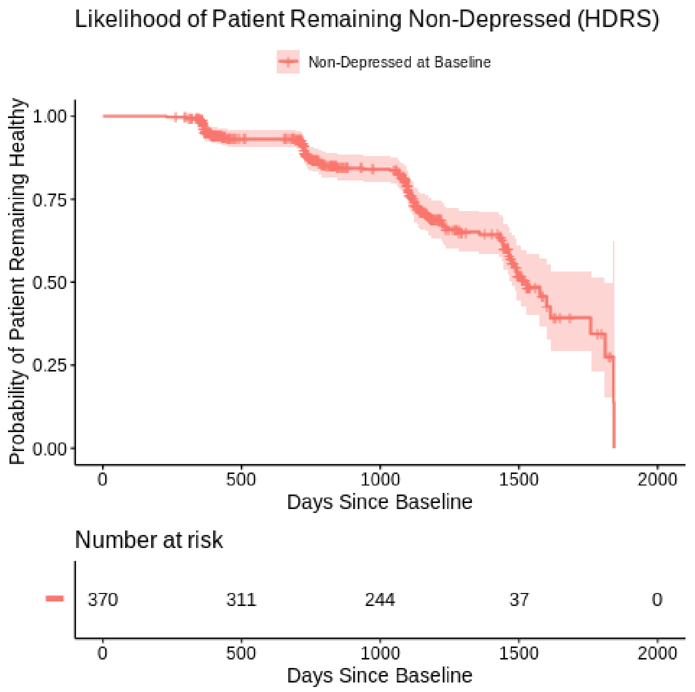


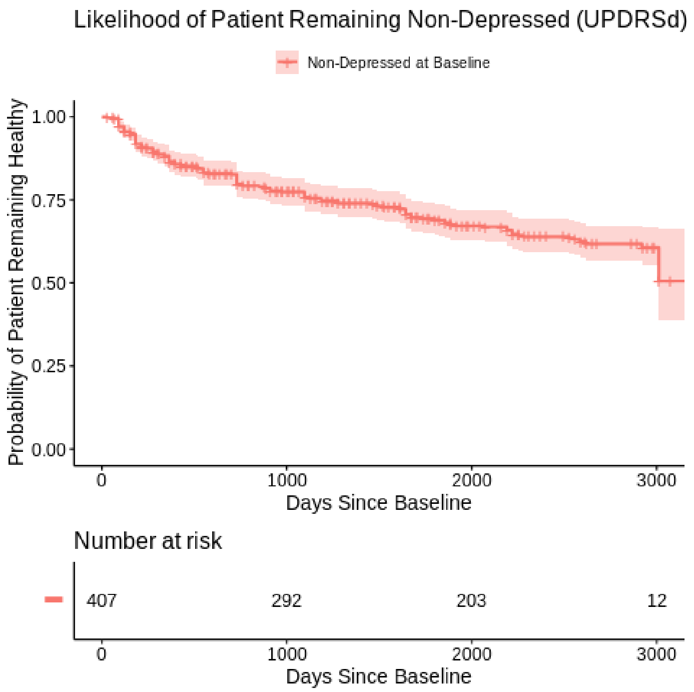


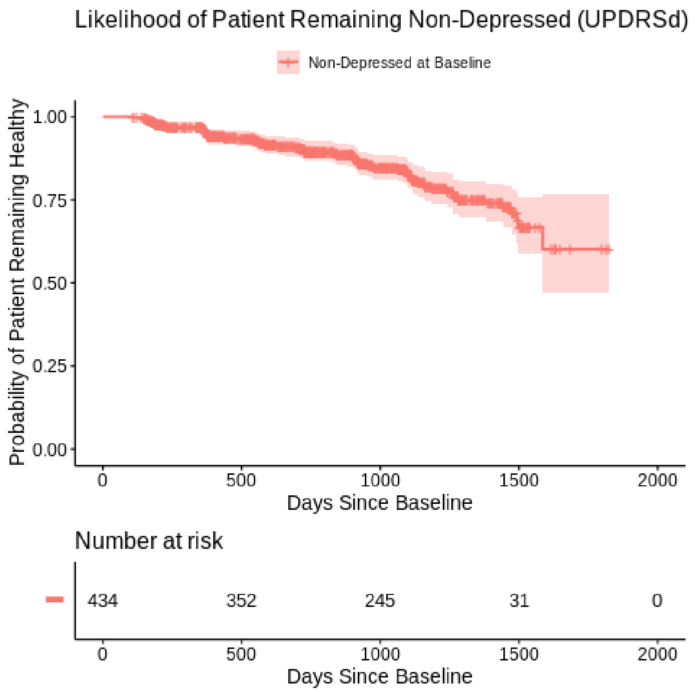


GDS, Geriatric Depression Scale; HDRS, Hamilton Depression Rating Scale; UPDRSd, Movement Disorder Society revised Unified Parkinson's Disease Rating Scale Part I Item 3

**Supplemental Table 1**
Results of linear mixed effects model analyzing the linear relationship between time since baseline and scores on various measures of depressive symptoms.

| PPMI | | | PDBP | | |
|---|---|---|---|---|---|
| Item [score range] | Estimate [95%C.I.] | p-value | Item [score range] | Estimate [95%C.I.] | p-value |
| GDS Total Score [0,15] | 0.086 [0.067, 0.106] | <2e-16 | HDRS Total Score [0,31] | 0.679 [0.56, 0.80] | <2e-16 |
| UPDRSd [0, 4] | 0.026 [0.018, 0.034] | 2.0e-9 | UPDRSd [0,4] | 0.068 [0.049, 0.087] | 3.1e-12 |

GDS, Geriatric Depression Scale; HDRS, Hamilton Depression Rating Scale; UPDRSd, Movement Disorder Society revised Unified Parkinson's Disease Rating Scale Part I Item 3

**Supplemental Figure 2**

PPMI Cohort (with baseline adjusted to account for difference in median duration of diagnosis)

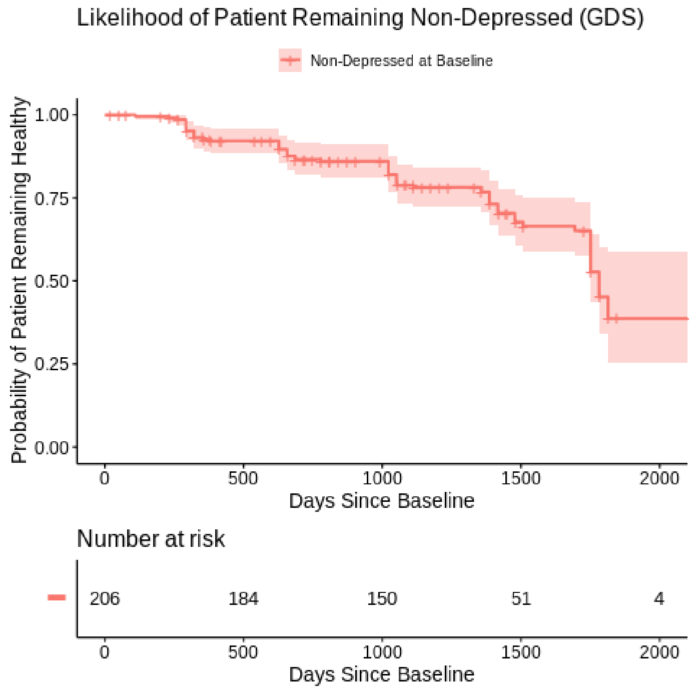


PDBP (with unchanged baseline)

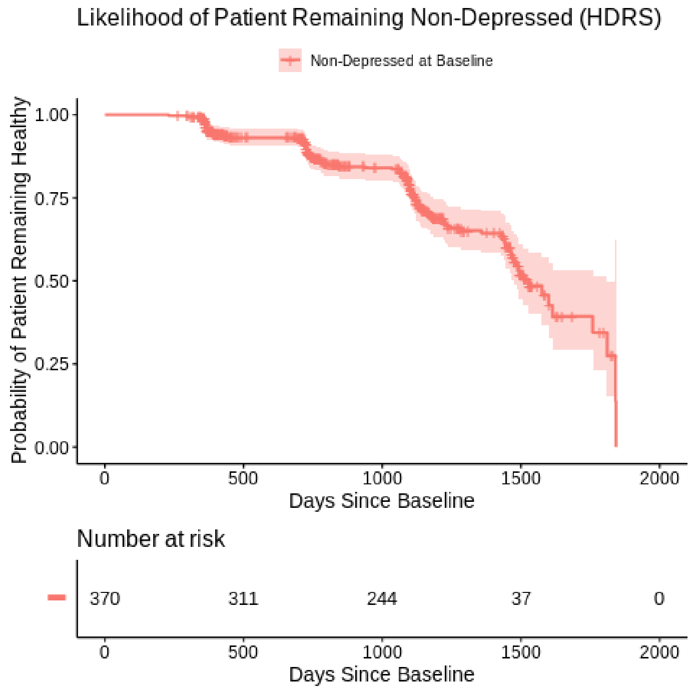


GDS, Geriatric Depression Scale; HDRS, Hamilton Depression Rating Scale

**Supplemental Figure 3**
Kaplan-Meier curves with number at risk tables for variables that were associated with an increased risk of depression in either cohort using their respective depression-specific scales.

| **PPMI** | **PDBP** |
|---|---|

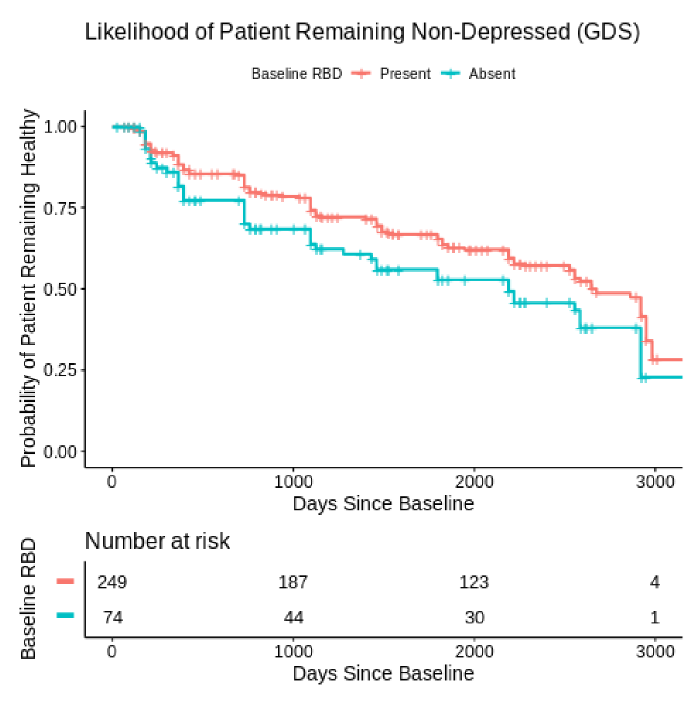


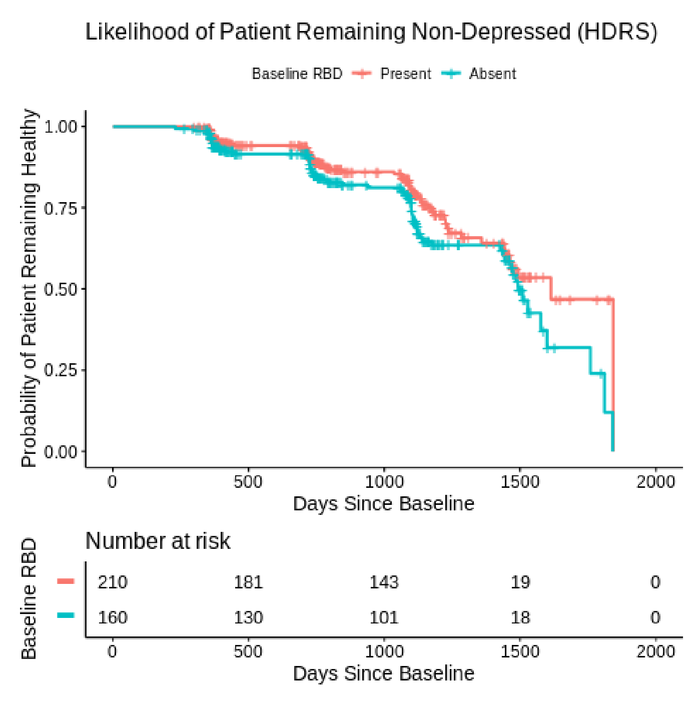


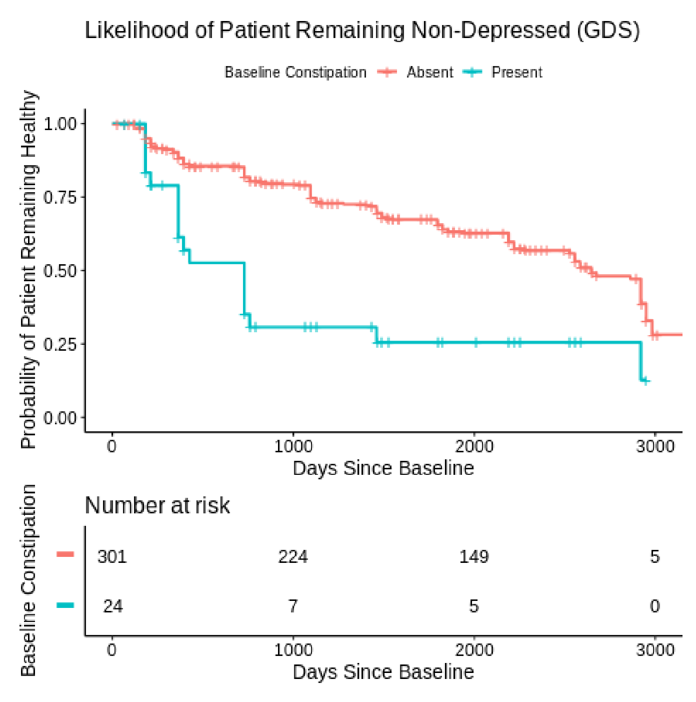


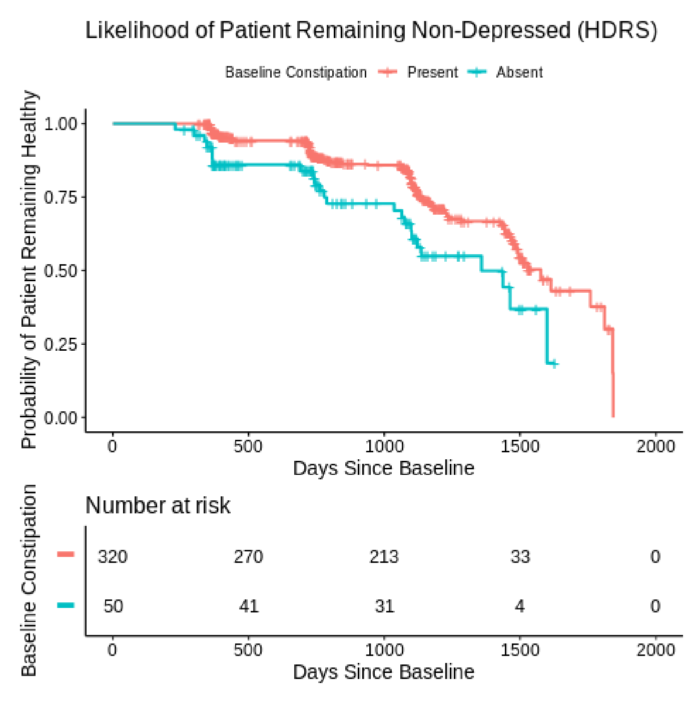

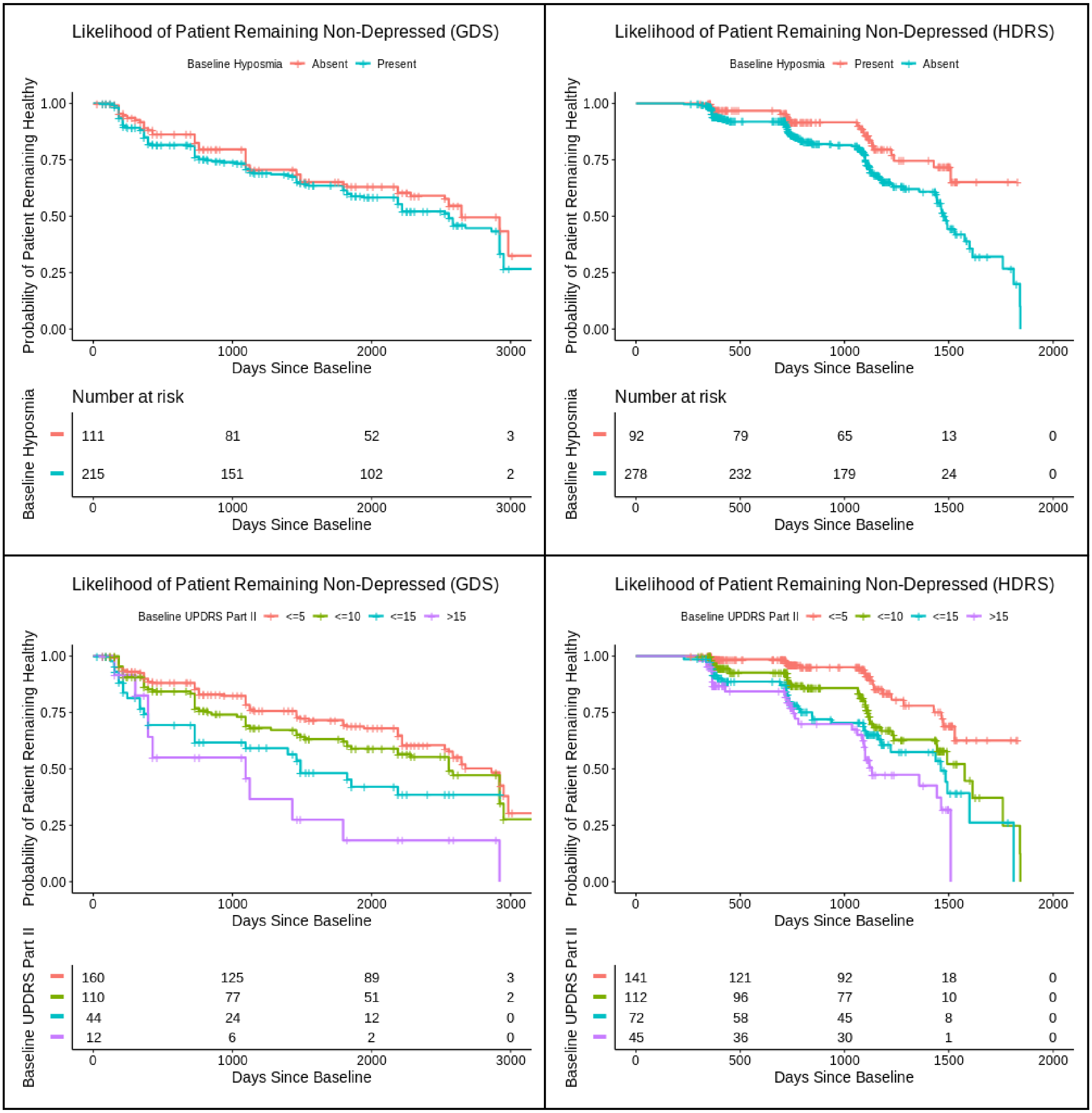
Likelihood of Patient Remaining Non-Depressed (GDS)
Baseline Hyposmia Absent Present
Probability of Patient Remaining Healthy
1.00
0.75
0.50
0.25
0.00
0
1000
2000
3000
Days Since Baseline
Number at risk
Baseline Hyposmia
111 81 52 3
215 151 102 2
0 1000 2000 3000
Days Since Baseline
Likelihood of Patient Remaining Non-Depressed (HDRS)
Baseline Hyposmia Present Absent
Probability of Patient Remaining Healthy
1.00
0.75
0.50
0.25
0.00
0
500
1000
1500
2000
Days Since Baseline
Number at risk
Baseline Hyposmia
92 79 65 13 0
278 232 179 24 0
0 500 1000 1500 2000
Days Since Baseline
Likelihood of Patient Remaining Non-Depressed (GDS)
Baseline UPDRS Part II <=5 <=10 <=15 >15
Probability of Patient Remaining Healthy
1.00
0.75
0.50
0.25
0.00
0
1000
2000
3000
Days Since Baseline
Baseline UPDRS Part II
160 125 89 3
110 77 51 2
44 24 12 0
12 6 2 0
0 1000 2000 3000
Days Since Baseline
Likelihood of Patient Remaining Non-Depressed (HDRS)
Baseline UPDRS Part II <=5 <=10 <=15 >15
Probability of Patient Remaining Healthy
1.00
0.75
0.50
0.25
0.00
0
500
1000
1500
2000
Days Since Baseline
Baseline UPDRS Part II
141 121 92 18 0
112 96 77 10 0
72 58 45 8 0
45 36 30 1 0
0 500 1000 1500 2000
Days Since Baseline

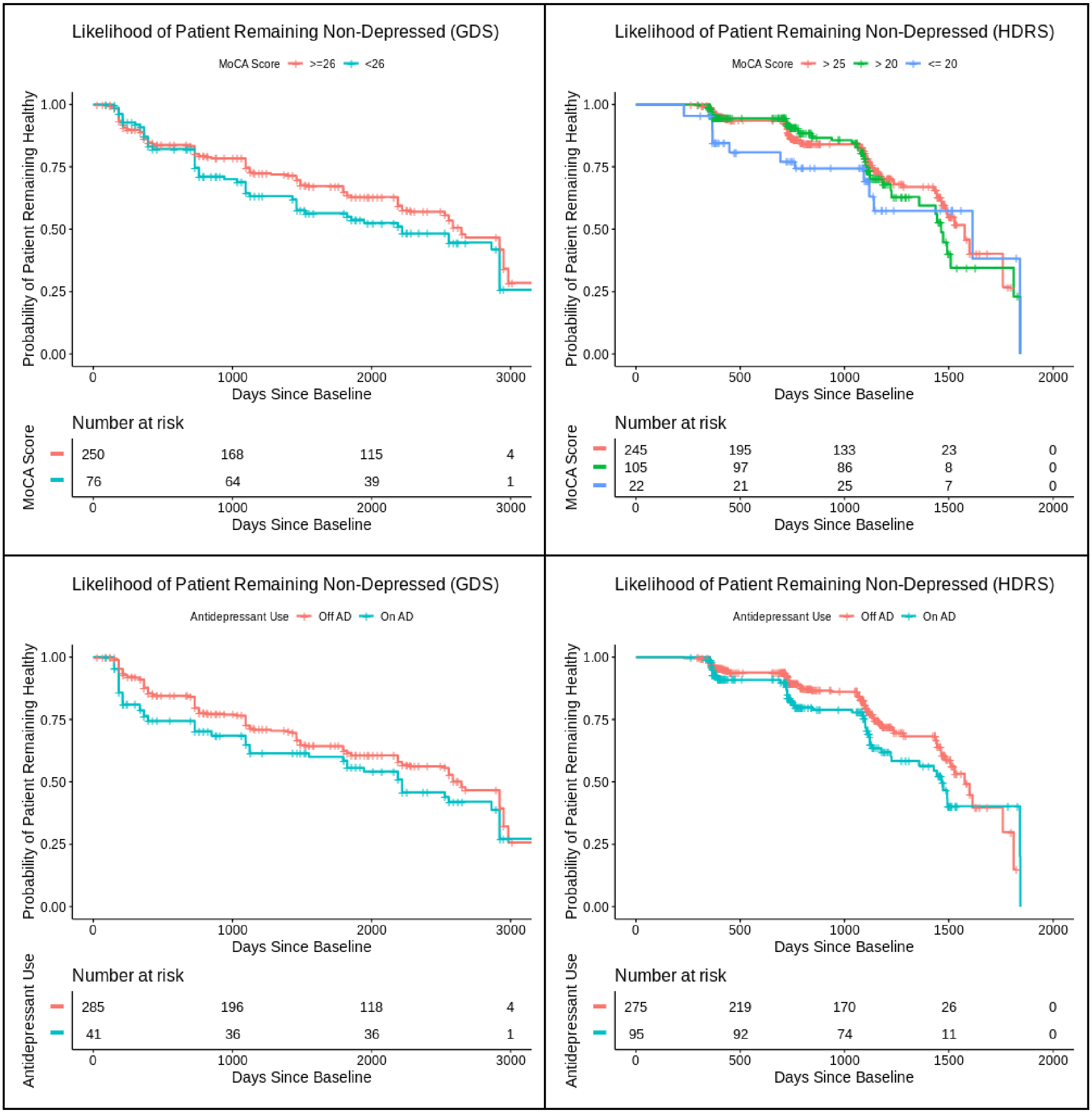

Likelihood of Patient Remaining Non-Depressed (GDS)
MoCA Score >=26 <26
Probability of Patient Remaining Healthy
1.00
0.75
0.50
0.25
0.00
0
1000
2000
3000
Days Since Baseline
Number at risk
MoCA Score
250 168 115 4
76 64 39 1
Likelihood of Patient Remaining Non-Depressed (HDRS)
MoCA Score > 25 > 20 <= 20
Probability of Patient Remaining Healthy
0
500
1000
1500
2000
Days Since Baseline
Number at risk
MoCA Score
245 195 133 23 0
105 97 86 8 0
22 21 25 7 0
Likelihood of Patient Remaining Non-Depressed (GDS)
Antidepressant Use Off AD On AD
Probability of Patient Remaining Healthy
Days Since Baseline
Number at risk
Antidepressant Use
285 196 118 4
41 36 36 1
Likelihood of Patient Remaining Non-Depressed (HDRS)
Antidepressant Use Off AD On AD
Probability of Patient Remaining Healthy
Days Since Baseline
Number at risk
Antidepressant Use
275 219 170 26 0
95 92 74 11 0

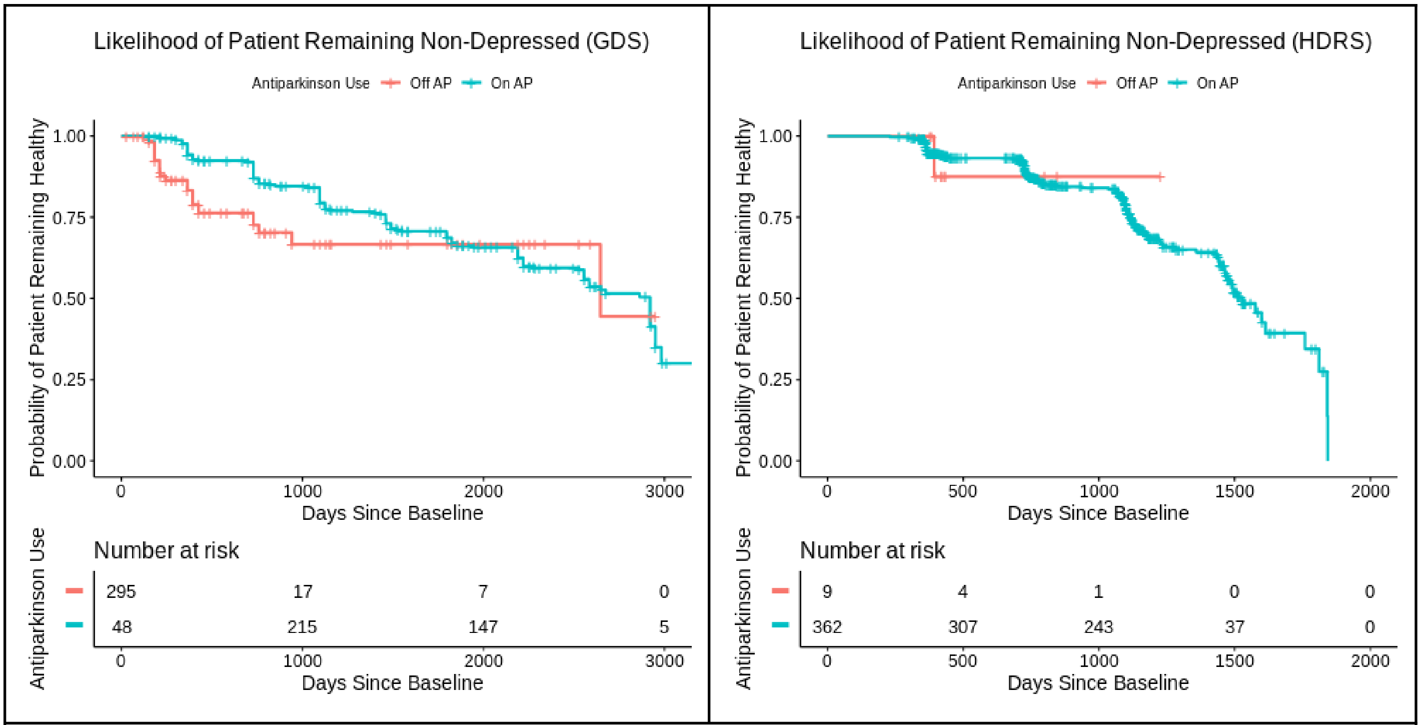


GDS, Geriatric Depression Scale; HDRS, Hamilton Depression Rating Scale; AD, Antidepressant; AP, Antiparkinson

**Supplemental Table 2**

Summary of meta-analysis of the cox proportional hazards models for depression in PPMI and PDBP using the UPDRS depressed mood item.

| | PPMI | | PDBP | | Meta-Analysis | | |
|---|---|---|---|---|---|---|---|
| | HR [95% CI] | p-value | HR [95% CI] | p-value | HR [95% CI] | p-value | I^2 |
| *Baseline Predictive Covariates (Time-Independent)* | | | | | | | |
| College Education or Greater | 0.85 [0.60, 1.22] | 0.38 | 0.62 [0.46, 0.85] | 2.7e-3 ** | 0.72 [0.53, 0.98] | 0.04 * | 42.6% |
| Gender (Male as Reference) | 1.46 [1.02, 2.08] | 0.04 * | 1.16 [0.90, 1.50] | 0.26 | 1.25 [1.01, 1.55] | 0.04* | 3.5% |
| Age at Baseline | 1.01 [0.99, 1.03] | 0.45 | 0.98 [0.95, 1.01] | 0.16 | 1.00 [0.97, 1.02] | 0.74 | 61.2% |
| Baseline Duration of Diagnosis | 0.83 [0.60, 1.15] | 0.26 | 1.00 [0.95, 1.06] | 0.97 | 0.98 [0.86, 1.11] | 0.71 | 14.6% |
| Baseline UPDRS Part II Total | 1.05 [1.01, 1.10] | 0.02 * | 1.06 [1.04, 1.09] | 3.6e-7 *** | 1.06 [1.04, 1.08] | <1e-4 *** | 0.0% |
| Probable RBD at Baseline | 1.19 [0.81, 1.75] | 0.39 | 1.49 [1.22, 1.81] | 7.1e-5 *** | 1.42 [1.18, 1.70] | 2e-4 *** | 3.19% |
| Baseline Hyposmia | 1.22 [0.84, 1.76] | 0.31 | 1.17 [0.79, 1.74] | 0.43 | 1.19 [0.91, 1.57] | 0.20 | 67.2% |
| Baseline Presence of Constipation | 2.74 [1.63, 4.59] | 1.3e-4 *** | 1.32 [1.02, 1.71] | 0.04 * | 1.83 [0.90, 3.74] | 0.10 | 83.7% * |
| Adjustment Covariates (Time-Dependent) | | | | | | | |
| MoCA Total Score | 0.97 [0.91, 1.03] | 0.29 | 0.99 [0.94, 1.04] | 0.77 | 0.98 [0.95, 1.02] | 0.36 | 0.0% |
| Use of Antidepressants | 2.62 [1.81, 3.80] | 3.7e-7 *** | 1.99 [1.06, 3.73] | 0.03 * | 2.44 [1.77, 3.36] | <1.0e-4 *** | 0.0% |

RBD, REM Sleep Behavior Disorder. UPDRS, Movement Disorder Society revised Unified Parkinson's Disease Rating Scale; MoCA, Montreal Cognitive Assessment;
***: p< 0.001, **: p<0.01, *: p<0.05. (t-test).
